# Raising the alarm: Endotoxin contamination in lithium heparin tubes is a threat to cell-based assay reliability

**DOI:** 10.64898/2025.12.18.25341698

**Authors:** Chrystelle Briere, Nathalie Vallon, Sandrine Ducrot, Camille Pease, Soizic Daniel

## Abstract

**Objective:** *In vitro* cytokine release assays are essential tools in immunological research, yet their reliability can be influenced by various pre-analytical factors. This study was initiated to investigate the potential impact of lithium heparin whole blood collection tubes from different manufacturers on assay performance. The objective of this work is to urge the scientific community to consider whole blood collection tubes as a significant and often underestimated source of variability in cell-based assays.

**Results description:** 210 whole blood samples from healthy donors were collected in plastic lithium heparin tubes sourced from 20 different suppliers worldwide. Testing revealed that 8 out of 20 of these tubes contained endotoxin levels >0.1□EU/mL, which triggered significant variability in cytokine responses (IFN-γ and IL-6) in cell-based assays. A dose-response curve using LPS from *E. coli* confirmed the impact of endotoxins on interferon-γ production.

## Introduction

Pre-analytical factors play a critical role in the reliability of cell-based assays, particularly those assessing immune or inflammatory responses. Among these, the presence of endotoxins-lipopolysaccharide (LPS)-derived contaminants in whole blood collection tubes represents a potential yet often overlooked source of variability [1]. This research note aims to raise awareness about the heterogeneity in endotoxin levels found in lithium heparin blood collection tubes from different manufacturers. Even trace amounts of endotoxins can significantly influence cellular responses, highlighting the importance of careful selection and evaluation of whole blood collection tubes in experimental setups involving cell-based assays [2–4].

Data were generated by bioMerieux R&D immunoassay department in Marcy l’Etoile, France through testing of 210 whole blood samples collected from EFS blood donors in lithium heparin tubes between September and December 2024. The objective was to characterize the extent of endotoxin contamination across different lithium heparin tube manufacturers and to investigate the relationship between endotoxin concentration and impact on an interferon gamma (IFN-γ) secretion cell-based assay.

## Main text

In this study, twenty different lithium heparin (LH) blood collection tubes from various manufacturers (mostly from Asia and Europe) were evaluated against a reference tube which has been used consistently in our laboratory. For each tube, whole blood (4-5mL) was collected from 10 healthy blood donors (age range: 18 to 70) in a randomized sequence between the reference and test tubes. Each patient sample was collected in a tube from a “test” tube as well as in a “reference” tube. Samples were transported under controlled conditions (2–8□°C) from the donor site to the testing laboratory within 8 hours and processed on the same day.

Whole blood samples were subsequently diluted 1:1 with phosphate-buffered saline (PBS) and incubated at 37□°C for 16 hours prior to testing. Interferon-γ (IFN-γ) concentrations were measured using the VIDAS^®^ platform with a sandwich ELFA Immunoassay (Enzyme Link Fluorescence Assay). The Relative Fluorescence Value (RFV) obtained is calculated as the difference between baseline fluorescence (measured before substrate incubation) and fluorescence measured after substrate incubation. This value represents the enzymatic activity corrected for background noise. The value of the fluorescence signal is proportional to the concentration of IFN-γ. The RFV signal from each patient sample was compared between the “test” tube and the “reference” tube, providing a ratio of secreted IFN-γ. For the “reference” tube, the ratio of secreted IFN-γ was done across 2 different manufacturer lots of reference supplier.

Additionally, endotoxin concentrations in each collection tube were measured using a recombinant factor C (rFc) endotoxin assay (endozyme^®^ II GO, bioMerieux), following manufacturer’s instruction for use. The reference tube was tested negative for endotoxins across several lots. Lithium heparin tubes were found to contain variable levels of endotoxin contamination. This contamination can trigger non-specific stimulation of immune cells, leading to inaccurate cytokine measurements [5] such as interferon-γ (IFN-γ) (Figure□1) or IL-6 (data not shown).

**Fig 1.**
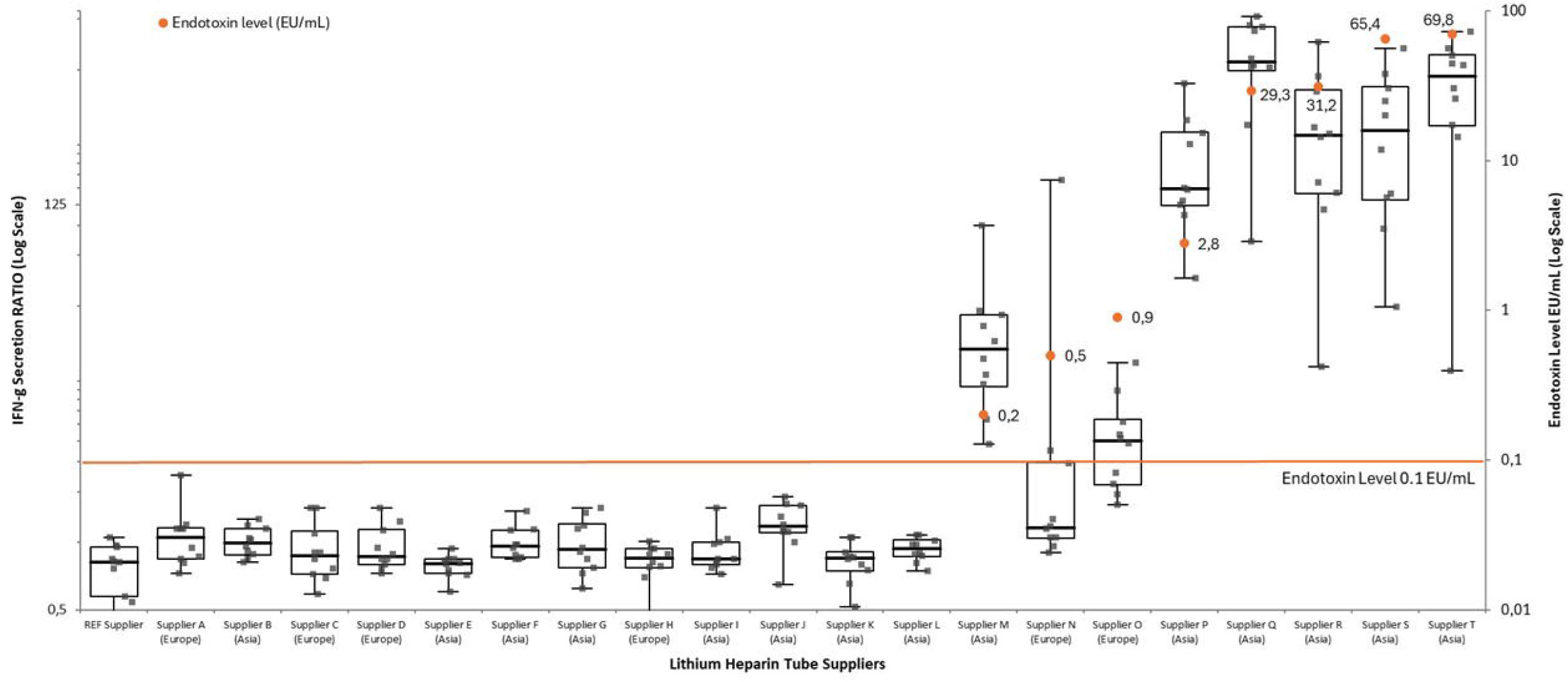

To assess tube performance under varying endotoxin loads, a dose–response curve was generated by spiking tubes with endotoxins (LPS from E.*coli* O113:H10) at different concentrations, and each level was tested using a subset of patient samples.

In conclusion, endotoxin contamination in lithium heparin tubes is a threat to the reliability of cell-based assays. It is recommended that blood collection tubes be tested for endotoxins before use. Also, it is theoretically possible that cell-based assays are sensitive to other additives or components of plastic tubes, other than bacterial contamination. We therefore recommend that collection tubes undergo formal validation to verify their compatibility with the intended application, in accordance with CLSI GP34 guidelines. Particular attention should be given to cell-based assays, where preanalytical variability can critically affect results. Moreover, strict adherence to good laboratory practices during tube handling is essential to minimize the risk of contamination of collection tube and ensure reliable performance.

## Limitations

This study focused on assessing the impact and variability in endotoxin levels across a broad range of lithium heparin blood collection tubes from different manufacturers. Only a single lot per supplier was evaluated, which limits the ability to draw conclusions about the reproducibility or consistency of endotoxin contamination. Additionally, to isolate the potential impact of endotoxins on assay specificity, particularly in cytokine release assays, only healthy blood donors were included. Further studies are needed to explore the effects of endotoxin contamination in samples from patients with underlying diseases or comorbidities.

We did not assess potential differences in IFN-γ release induced by endotoxins originating from various bacterial strains that could contaminate LH tubes. Our dose–response experiments were restricted to endotoxins derived from *E. coli*, which may not fully represent the effects of endotoxins from other bacterial sources. As a result, the relationship between endotoxin quantity and cellular stimulation cannot be fully extrapolated.

Furthermore, this study focused exclusively on a single tube type, 4–5□mL plastic lithium heparin vacuum tubes, which represent the reference tube currently employed in our laboratory. Consequently, we did not evaluate whether other tube components, such as stoppers, lubricants, or anticoagulants, might influence IFN-γ secretion.

Finally, the documentation provided by blood collection tube manufacturers did not consistently include sufficient detail regarding production processes and material composition. This limited our ability to systematically compare specific manufacturing parameters, such as sterilization methods, to determine which tube references might be less prone to endotoxin contamination and the underlying reasons for these differences.

## Abbreviations

LPS: lipopolysaccharide
IFN-γ: interferon-gamma
IL-6: interleukin-6
RFV: Relative Fluorescence Value.

## Declarations

### Ethics approval and consent to participate

This research had been performed in accordance with the Declaration of Helsinki. Human samples used were residual volumes of whole blood obtained during routine blood donation procedure from the French Blood Agency (EFS Auvergne Rhône Alpes, France). Donors were duly informed and provided consent for the reuse of their samples for research purposes, approved by Ministry of Higher Education and Research (authorization number CODECOH AC-2020-3959 for EFS and CODECOH DC-2024-6831 for bioMerieux). In accordance with Article L1211-2 of the French Public Health Code and the provisions of the Jarde Law, no additional consultation with a Comite de Protection des Personnes (CPP) was required. Participation was on a voluntary basis; data security and anonymity were guaranteed (in accordance with French regulations MR-003).

## Consent for publication

Not applicable

### Data availability

The raw data supporting the findings of this study are not publicly available in the first instance but can be provided upon reasonable request.

### Funding

This research was funded by bioMérieux.

### Authors’ contributions

C.Briere contributed to the conception of the study,

S. Daniel and C. Pease wrote the main manuscript text,

N. Vallon and S. Ducrot contributed to the acquisition and analysis of generated data,

All authors reviewed the manuscript.

## Acknowledgments

We thank the Etablissement Français du Sang, National French blood bank EFS Auvergne-Rhone-Alpes, notably Yves Mérieux, for providing samples.

We thank Laurine Jacquet, Veronique Alberti, Sandrine Ducrot, Nathalie Vallon, Christian Faderl and Didier Poirault for their valuable contributions to the provision of patient samples and sample collection materials as well as for their support in the execution, and data analysis of this research.

## References

1. Jorda A, Eberl S, Nussbaumer-Pröll A, et al. Reproducibility of LPS-Induced ex vivo Cytokine Response of Healthy Volunteers Using a Whole Blood Assay. J Inflamm Res. 2024. doi:10.2147/JIR.s459999.

2. Newhall KJ, Diemer GS, Leshinsky N, Kerkof K, Chute HT, Russell CB, Rees W, Welcher AA, Patterson SD, Means GD. Evidence for endotoxin contamination in plastic Na+-heparin blood collection tube lots. Clin Chem. 2010. doi:10.1373/clinchem.2006.144618. Epub 2010 Jul 27. PMID: 20663962.

3. Aziz N, Irwin MR, Dickerson SS, Butch AW. Spurious tumor necrosis factor-alpha and interleukin-6 production by human monocytes from blood collected in endotoxincontaminated vacutainer blood collection tubes. Clin Chem. 2004. doi:10.1373/clinchem.2004.040162. PMID: 15502103.

4. Stoddard MB, Pinto V, Keiser PB, Zollinger W. Evaluation of a whole-blood cytokine release assay for use in measuring endotoxin activity of group B Neisseria meningitidis vaccines made from lipid A acylation mutants. Clin Vaccine Immunol. 2010. doi: 10.1128/CVI.00342-09. Epub 2009 Nov 18. PMID: 19923573; PMCID: PMC2812078.

5. Slater M, Dubose A, Banaei N. False-positive quantiferon results at a large healthcare institution. Clin Infect Dis. 2014. doi: 10.1093/cid/ciu139. Epub 2014 Mar 6. PMID: 24610428; PMCID: PMC4031620.

